# Recovery of Neurophysiological Measures in Post-COVID Fatigue – a 12-month Longitudinal Follow-up Study

**DOI:** 10.1101/2023.11.22.23298846

**Authors:** Natalie J. Maffitt, Maria Germann, Anne M.E. Baker, Mark R. Baker, Stuart N. Baker, Demetris S. Soteropoulos

## Abstract

One of the major consequences of the COVID-19 pandemic has been the significant incidence of persistent fatigue following resolution of an acute infection (i.e. post-COVID fatigue). We have shown previously that, in comparison to healthy controls, those suffering from post-COVID fatigue exhibit changes in muscle physiology, cortical circuitry, and autonomic function. Whether these changes preceded infection, potentially predisposing people to developing post-COVID fatigue, or whether the changes were a consequence of infection was unclear. Here we present results of a 12-month longitudinal study of 18 participants from the same cohort of post-COVID fatigue sufferers to investigate these correlates of fatigue over time. We report improvements in self-perception of fatigue via questionnaires, as well as significant improvements in objective measures of peripheral muscle fatigue and autonomic function, bringing them closer to healthy controls. Additionally, we found reductions in muscle twitch tension rise times, becoming faster than controls, suggesting that the improvement in muscle fatigability might be due to a process of adaptation rather than simply a return to baseline function.

## Introduction

In May 2023, more than three years into the pandemic, the World Health Organization (WHO) Emergency Committee on COVID-19 recommended that the pandemic no longer represented a public health emergency of international concern^1^. However, while acute COVID-19 may no longer constitute a global emergency, the long-term effects of severe acute respiratory syndrome coronavirus 2 (SARS-CoV-2) infection are now becoming apparent. Although the majority of infected individuals experience mild symptoms or recover within a few weeks, a significant proportion continue to experience persistent symptoms beyond the acute phase^2-4^, a condition commonly known as long COVID or post-acute sequelae of SARS-CoV-2 infection.

Long COVID is characterized by a diverse range of symptoms, including fatigue, cognitive impairment, respiratory issues, and musculoskeletal problems^5-7^. Among these, muscle pathology has emerged as a common and debilitating feature, even in individuals with a history of mild COVID-19^8^. Reports highlight muscle weakness, myalgia, and impaired motor coordination among long COVID patients^2,5,9^, significantly impacting their quality of life and functional abilities.

Physiological muscle stimulation during physical activity leads to the production of myokines, which normally induce an anti-inflammatory environment. However, in the presence of the SARS-CoV-2 virus, myokine production instead stimulates a prolonged muscular inflammatory environment^10,11^. Inflammation impairs muscle protein synthesis and mitochondrial activity, and thus has the potential to induce long-term sarcopenia^12^.

To form a comprehensive understanding of the mechanisms underlying long COVID, longitudinal studies are crucial. These allow the evaluation of clinical, immunological, radiological and neurophysiological changes over time, providing insights into the patterns of recovery of these metrics. Comparison of recovery time courses between clinical measures and biomarkers could potentially address whether abnormal measures play a causal role in the disease, or are simply correlated with its downstream consequences.

In a previous study^13^ we demonstrated changes in muscle physiology in long COVID after non-severe SARS-CoV-2 infection, where there had been no requirement for acute hospital in-patient care. These changes might explain some of the symptoms of fatigue described by patients. Here we report a longitudinal follow-up study in a subset of the same cohort, in whom neurophysiological measurements and subjective perception of fatigue were repeated twice more, at 6 month intervals. Levels of fatigue improved significantly after a year; for most of the neurophysiological metrics there was likewise a return to levels seen in age and sex-matched controls. We therefore suggest that these changes in muscle physiology and autonomic function occur as a consequence of a SARS-CoV-2 infection, rather than a pre-existing phenotype that increased susceptibility to developing post-COVID fatigue.

## Methods

Our previous publication^13^ employed 35 non-invasive behavioural and neurophysiological tests to assess specific circuits within the central, peripheral and autonomic nervous systems. In this longitudinal follow up study, we used only the sub-set of these tests which were significantly different between people with pCF and controls in our original paper. Transcranial Magnetic Stimulation (TMS) allowed us to probe the state of cortical motor circuits. Electrical stimulation of muscles assessed peripheral fatigue, recordings of heart rate assessed the state of the autonomic nervous system, and blood oxygen saturation (SaO_2_) was also collected. Participants completed a fatigue impact scale (FIS) questionnaire via a web-based survey tool.

### Participants

The study was approved by the Ethics Committee of Newcastle University Faculty of Medical Sciences; participants provided written informed consent.

Measures collected in our initial study^13^ were used here as baseline data. This included data from a cohort of 37 participants (27 female) who were suffering from pCF by self-report and a second cohort of 52 volunteer controls (37 female) with no symptoms of fatigue. Inclusion criteria were age 18–65 years, with no history of neurological disease. The first visit to the laboratory was made 6–26 weeks after infection for the pCF cohort. In the control cohort, six subjects had reported having mild COVID-19 but with complete recovery and no symptoms of pCF. Of the 37 people with pCF, 18 participated in this longitudinal follow-up study (13 female), completing a further two lab visits at intervals of approximately 6 months to yield a total of three visits.

### General Electrophysiological Methods

Electromyographic activity (EMG) was recorded with adhesive surface electrodes positioned over muscles (Kendall H59P, Covidien, Dublin, Ireland) using an isolated amplifier (D360, Digitimer, Welwyn Garden City, UK; gain 500, bandpass 30 Hz-2 kHz). Transcranial magnetic brain stimulation (TMS) was given with a Bistim 200^2^ stimulator and figure-of-eight coil (7 cm diameter for each winding; Magstim Company Limited, Whitland, UK), with the coil held tangential to the head at around 45° to the parasagittal plane, inducing current in the brain from posterior to anterior. Coil position relative to the head was maintained using a Brainsight neuronavigation system (Brainbox, Cardiff, UK). Stimulus timing was controlled by a Power1401 intelligent laboratory interface running Spike2 software (Cambridge Electronic Design, Cambridge, UK), which also sampled EMG and other task-related signals to hard disk (sampling rate 5 kSamples/s). All measurements were made on the self-reported dominant side. Offline analysis was performed with custom scripts written in the MATLAB programming environment.

### Paired-Pulse TMS

EMG was recorded from the first-dorsal interosseous (1DI); the TMS coil was moved to locate the hot spot for this muscle. The resting motor threshold (RMT) was determined, as the intensity required to generate MEPs of amplitude greater than 100 μV on 3/6 sweeps. The test stimulus intensity was set to generate MEP amplitudes of 1 mV, or to 1.2xRMT, whichever was lower. The conditioning stimulus intensity was 0.8xRMT. We then measured the responses to the test stimulus alone, and when preceded by the conditioning stimulus at intervals of 10 ms, corresponding to intracortical facilitation (ICF) (see Baker et al., 2023 Supplementary Figure 1). Twenty repetitions of each condition were given, in pseudo-random order, with the subject at rest. Offline analysis measured the peak-to-peak amplitude of responses to conditioned stimuli as a percentage of the responses to test stimulus alone, yielding the measure *TMS_ICF*.

### Heart Rate

A single channel electrocardiogram (ECG) recording was made, using a differential recording from either left shoulder and right leg, or left and right shoulders (bandpass 0.3-30 Hz, gain 500, sampling rate 1 kSamples/s). The ECG was processed offline to extract the time of each QRS complex and compute the mean heart rate (measure *Mean_HR*). Heart rate measures were made during a Stop Signal Reaction Time test, which ensured that the subject was sitting quietly, while engaged in a consistent behaviour.

### Measures of Muscle Physiology

The twitch interpolation procedure allows assessment of an individual’s ability to activate a muscle maximally voluntarily; in this study, we also measured changes after a sustained (fatiguing) contraction^14^. The protocol followed previous work from this laboratory^15^ and was identical to that used in Baker, et al. ^13^. Subjects sat with their dominant arm and forearm strapped into a dynamometer to measure torque about the elbow; the shoulder was flexed, and the elbow at a right angle, so that the upper arm was horizontal and the forearm vertical. The forearm was supinated. Thin stainless-steel plate electrodes (size 30x15 mm) were wrapped in saline-soaked cotton gauze and taped over the belly of the biceps muscle (cathode) and its distal tendon (anode). Electrical stimuli were delivered through these electrodes while monitoring the evoked twitch response recorded by the dynamometer, and the intensity increased until the response grew no further. This supramaximal stimulus was used for all subsequent measurements.

The following recordings were then made in sequence; this protocol was followed to maintain consistency with our original study. A brief tone cued the subject to produce and hold a maximal voluntary contraction; 2 s after the tone, a stimulus was given to the biceps, and 1 s later a second tone indicated that the subject should relax. Five seconds later, a biceps stimulus was given, followed by a further 55 s rest period. This sequence was repeated three times. A long tone then cued the subject to make a sustained maximal voluntary contraction. This was continued either for 90 s, or until the force exerted fell to 60% of the initial maximal level. During this sustained contraction, the biceps was stimulated every 10 s. After the contraction ended, a final three biceps stimuli were given at rest (inter-stimulus interval 5 s).

From the three stimuli delivered at rest at the start, we averaged the maximal twitch at rest and measured its amplitude,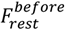. From the three stimuli delivered at rest after the sustained contraction, we measured 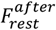.

Peripheral fatigue (measure *TI_PeriphFatigue*) was calculated as:

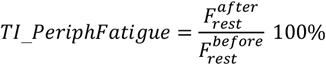

This describes the reduced ability of the muscle to generate force after fatigue, even when activation is performed independent of the central nervous system by an electrical stimulus to the muscle. Additionally, we measured the time to maximal force generation following direct electrical stimulation to the muscle (measure *Rise Time*) using the twitch initially evoked at rest.

### Biometric Data

Blood oxygen saturation was measured using a pulse oximeter placed onto the index finger (*SaO*_*2*_).

### Statistics

Descriptive statistics are given as mean ± standard deviation (SD). Comparisons for each metric across visits were carried out using a repeated measures ANOVA (using the ‘fitrm’ function in MATLAB). Correlation between measures was assessed with linear regression and results reported as r^2^ values. Paired t-tests were used to compare individual measures between visits. Unpaired t-tests were used to compare measures between different cohorts.

## Results

Figure 1A shows the time line of the repeat visits of the 18 participants (13 females; age 48.1±10.0 years) for whom we collected longitudinal data. Visits 2 (V2) and 3 (V3) were on average 6.02±0.34 and 11.61±0.36 months after visit 1 (V1).

**Figure 1.**
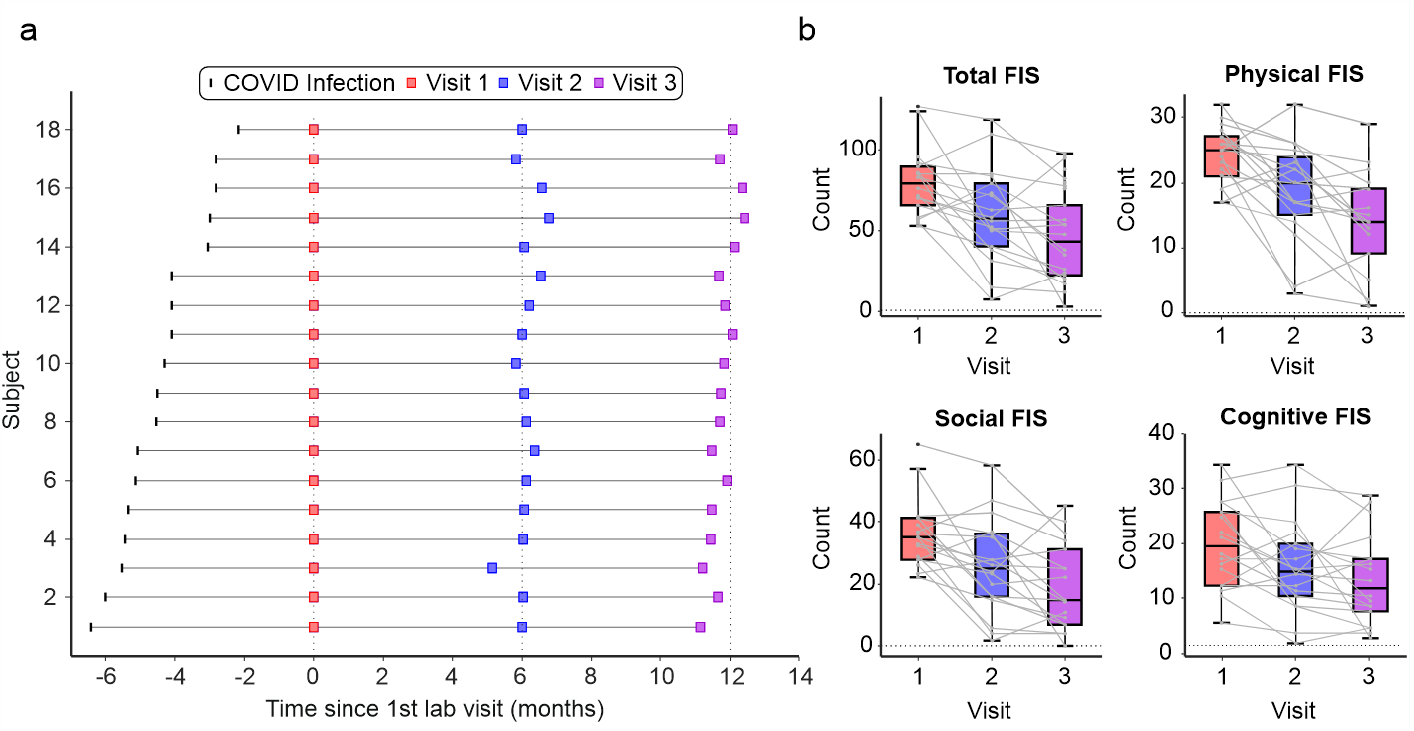
Timeline of visits and FIS scores. a) Timeline of SARS-CoV-2 infection and subsequent visits to the lab for assessment for each participant. b) Averaged FIS scores as box plots across all 18 participants for all visits; grey lines indicate individual subjects. Total FIS maximum count = 160; Physical = 40; Cognitive = 40; Social = 80.

### Fatigue Impact Scale

There was a significant (p<0.0001, F=13.3) decrease in the self-reported perception of fatigue across visits; mean FIS score declined from 80.3 at V1 to 60.2 at V2 and 46.1 at V3 (Figure 1B). A similar trend was seen for all of the sub-domains of the FIS score: there was a significant average decrease over time in the cognitive FIS score (by 6.8 from V1 to V3, p=0.009, F=5.5), the social FIS score (decrease 16.9, p<0.0001, F=14.00) and the physical FIS score (decrease 10.5, p<0.0001, F=19.14). Overall, the majority (16/18) of participants had improved FIS scores between V1 and V3.

### Changes in Biological Measures

Our earlier work showed that only a small number of measures (*TI_PeriphFatigue, TMS_ICF, Mean_HR, SaO*_*2*_) out of an extensive initial set were significantly different between controls and participants suffering from pCF. Only these measures were therefore repeated during V2 and V3 (Figure 2). Over time there was a significant change in peripheral oxygen saturation (*SaO*_*2*_, p=0.022, F=4.74), heart rate (*Mean_HR*, p=0.033, F=3.88), and peripheral fatigue (*TI_PeriphFatigue*, p=0.006, F=6.1). Using TMS to investigate the excitability of the primary motor cortex, we found that although intracortical facilitation became more similar to controls with each visit, this trend did not reach significance (*TMS_ICF*, p = 0.075, F=2.79).

**Figure 2.**
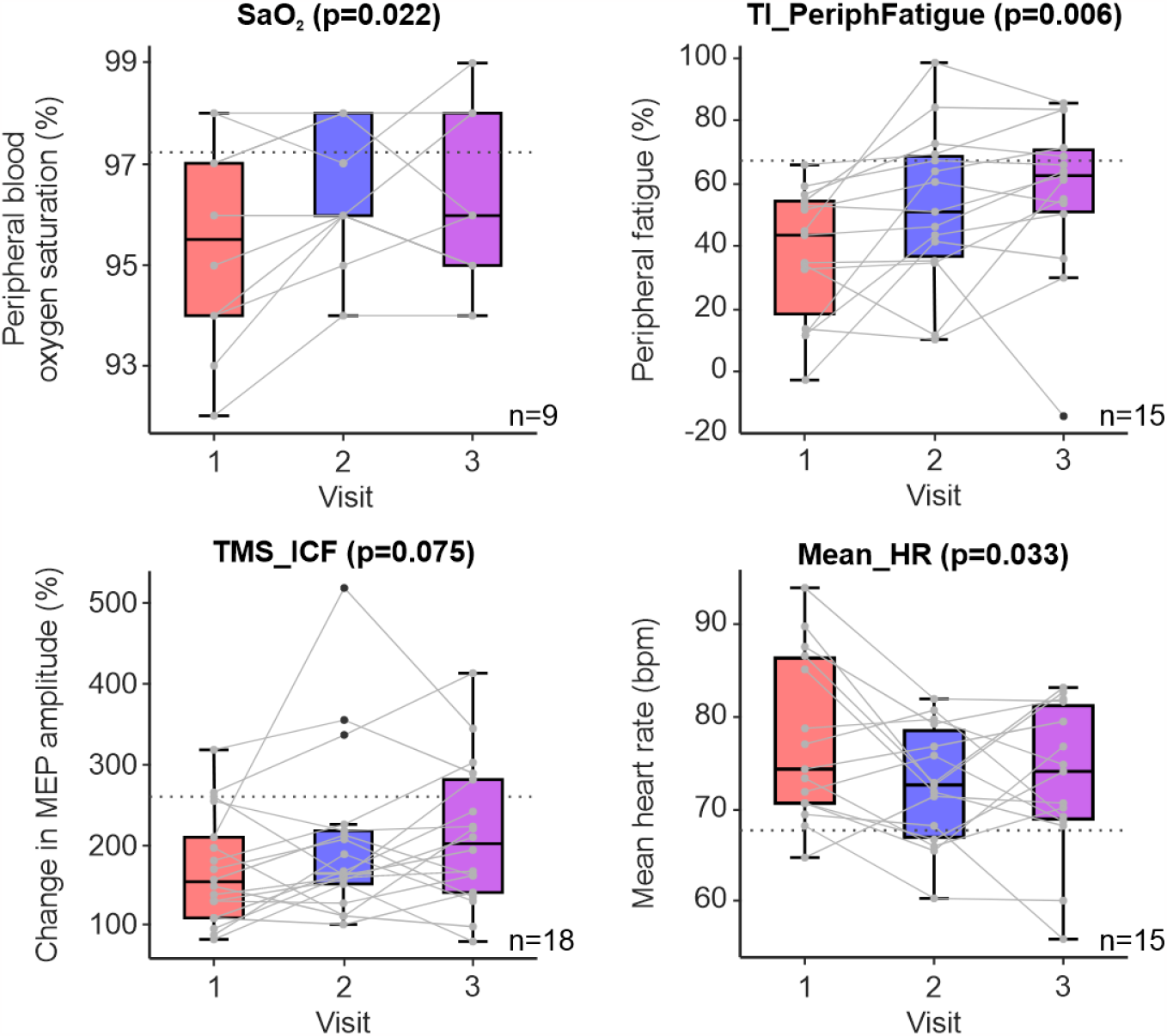
Changes in metrics over time. Box plot illustration of each metric (*SaO*_*2*_, *TI_PeriphFatigue, TMS_ICF, Mean_HR*) over time. The mean of the control cohort for each metric is illustrated by grey dotted lines. P values above each plot are taken from repeated measure ANOVAs for each metric.

Post hoc comparisons showed that *SaO*_*2*_ at V2 and V3 significantly increased from V1 (+0.71%, p=0.023 and +1.04%, p=0.015 respectively) but was not significantly different between V2 and V3. Similarly, *mean_HR* at V2 and V3 significantly decreased relative to V1 (-4.63 bpm, p=0.024 and - 5.78 bpm, p=0.035 respectively). *Mean_HR* was not significantly different between V2 and V3.

Peripheral fatigue (*TI_PeriphFatigue*) also showed significant improvements over time. At both V2 and V3, *TI_PeriphFatigue* significantly increased (indicating less peripheral fatigue) compared to V1 (+17.49%, p=0.01 and +18.02%, p=0.008 respectively), but values at V3 were not significantly different from those at V2.

Although both FIS scores and the biological metrics improved significantly over the course of the 12 month study, surprisingly there was no significant correlation between change in FIS scores and changes in any of these measures (r^2^ values: *TMS_ICF* 0.00063, *SaO*_*2*_ 0.011, *Mean_HR* 0.17, *TI_PeriphFatigue* 0.086, all p-values > 0.05).

### Additional Measures of Muscle Physiology

To investigate the potential mechanisms underlying improvements seen with peripheral fatigue over time, we made one further measure of muscle physiology - the rise time of a maximal twitch (*RiseTime*). This refers to the time taken from direct muscle stimulation to the peak force generated (measured from the biceps, at rest at the start of the twitch interpolation protocol; Figure 3A). Although, relative to controls, there was no significant difference at V1 (p=0.734), *RiseTime* did become significantly different from controls at both V2 (p=0.012) and V3 (p=0.002). Furthermore, *RiseTime* of the pCF cohort significantly changed over time (p<0.0001, F= 19.94). Post hoc comparisons showed that at both V2 and V3, *RiseTime* significantly decreased relative to V1 (-9.8 ms, p<0.001 and -11.8 ms, p<0.001 respectively). *RiseTime* was not significantly different between V2 and V3.

**Figure 3.**
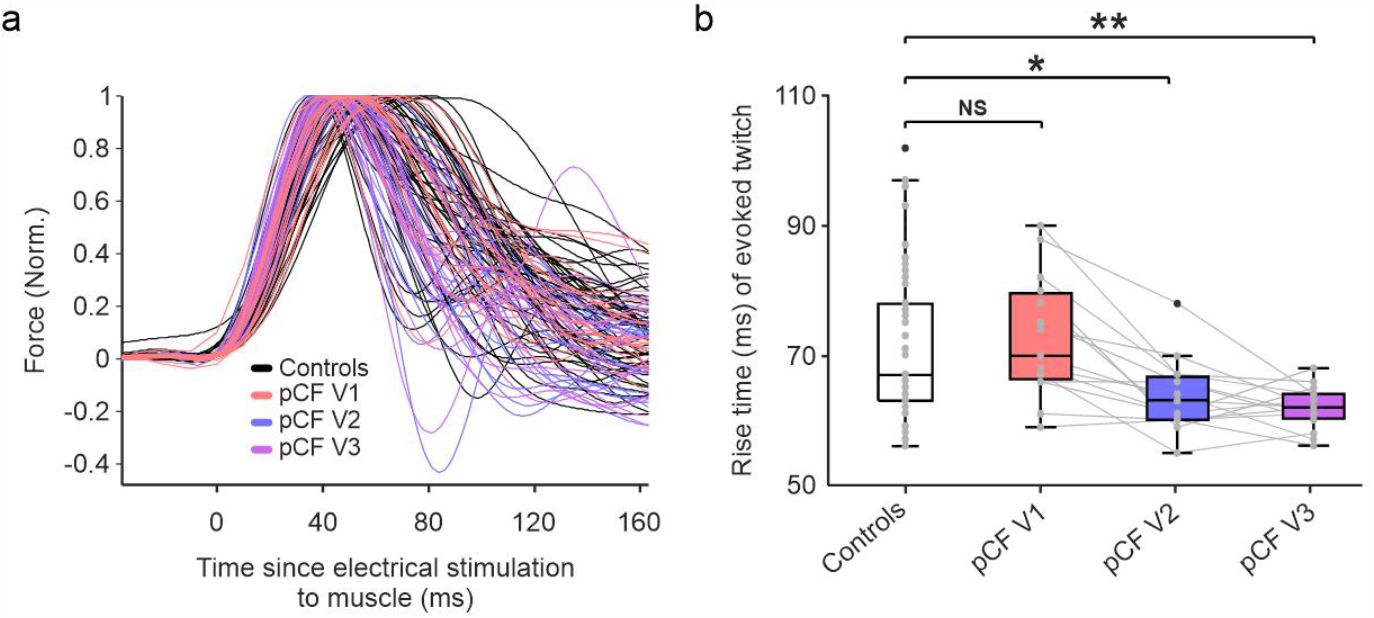
Rise Time. a) Raw traces of twitch responses to direct electrical stimulation of biceps for each participant, for all cohorts. The trace for each participant was normalised to the maximal force generated by each individual. *RiseTime* is calculated as time from electrical stimulation to maximal force generated. b) Box plots showing *RiseTime* data for controls (n=51) and post-COVID fatigue participants (n=15) for each visit. NS, not significant; * P<0.05; ** P<0.005.

## Discussion

A substantial proportion of people in the UK (2.9%) continue to suffer from longer-term sequalae of SARS-COV-2 infection (Long COVID)^16^. Of various lingering symptoms such as difficulty concentrating (51%), muscle aches (49%) and shortness of breath (48%), persistent fatigue is one of the most common (72%)^16^. Persistent fatigue after a (non-SARS-CoV-2) viral infection is well known to the clinician but is also a hallmark of several autoimmune and neurological disorders, suggesting a link with nervous system dysfunction. Recent work has shown that even a mild to moderate COVID-19 infection can cause dysregulation in the nervous system^17^.

In our previous study^13^, we provided evidence that measures of the central (intracortical facilitation, *TMS_ICF*), peripheral (peripheral fatigue, *TI_PeriphFatigue*) and autonomic (mean heart rate, *Mean_HR* and peripheral oxygen saturation, *SaO*_*2*_) nervous systems were different between people with pCF compared to sex- and age-matched controls. However, these findings could not infer causation; measures could have been different prior to the infection (and thus conferred an increased likelihood of developing pCF) or the abnormal measures could be a consequence of the infection (and hence might be useful as a biomarker for tracking changes in fatigue). To address this, we repeated measurements 6 months and 12 months later. As a group the reported levels of fatigue improved, and our metrics returned to or were returning to normal. This suggests that the changes were mediated by SARS-CoV-2 infection, rather than being a pre-existing long-term trait amongst the pCF sufferers.

The most significant change over time was observed in muscle fatigue. *TI_PeriphFatigue* increased from 37% at the first lab visit to 55% one year later, indicating that muscles undergo significant physiological changes during recovery. Acute SARS-COV-2 infection can affect skeletal muscle through several mechanisms. Firstly, the virus can cause direct muscle cell damage by entering host cells via the ACE-2 receptor and TMPRRSS2 protein^18^, both of which are expressed by musculoskeletal tissue^19^. In muscle, there is some evidence that SARS-CoV-2 may impair mitochondrial function directly causing critical illness myopathy^20^. Secondly, patients can develop acute respiratory distress syndrome^21,22^, characterised by severe hypoxaemia, thereby diminishing systemic oxygen supply to muscle tissue. Hypoxia significantly affects mitochondrial activity, compromising muscle energy generation needed for protein synthesis and muscle contraction^23,24^. Thirdly, SARS-COV-2 infection can cause a hyper-inflammatory state in muscle^10,11^. Inflammation, similar to hypoxia, also impairs mitochondrial function^25^.

Aside from the potential direct actions of SARS-CoV-2 on mitochondria (see above), there may also be indirect effects of SARS-COV-2 on mitochondrial function that contribute to ongoing fatigue encountered in long COVID. For example, there is evidence in critical illness that mitochondrial haplotype conveys survival advantage^26^. In sepsis, mitochondrial dysfunction results in diaphragmatic myopathy^27^, and the extent of mitochondrial DNA depletion, measurable in mononuclear cells, correlates with the severity of critical illness^28^ It is thus possible that those who develop pCF might have subclinical mitochondrial dysfunction, subsequently unmasked by SARS-COV-2 infection, leaving them with systemically impaired mitochondrial function, as is typical of the pattern in mitochondrial disease^29^, potentially explaining the broad spectrum of symptoms experienced by patients suffering from long COVID.

Whilst sepsis-related multi-system mitochondrial dysfunction may play a role in the pathophysiology of long COVID, there is clear evidence for a specific role of mitochondrial dysfunction in critical illness myopathy^30,31^. Presumably this is a result of metabolic adaptations, including a shift away from energy generation in mitochondria through oxidative phosphorylation and towards anaerobic glycolysis, where pyruvate is converted into lactate^32^, an excess of which favours the maintenance of inflammatory processes^33^.

Our cohort with pCF showed significantly less peripheral fatigue 6 months and 12 months after they were first assessed. Interestingly, as the pCF cohort recovered from the symptoms of fatigue, their twitch tension rise times became faster than controls (Figure 3B). This suggests that the improvement in fatigability of muscles over time might be due partly to a process of adaptation rather than a complete return to baseline physiology. We do not know whether rise time would eventually return to baseline values over a time course of more than the one year interval in our study.

Persistent muscle pathology, initiated in the acute phase of the disease and perhaps best explained by mitochondrial dysfunction, could lead to dysfunction in type 1 muscle fibres. Although this is speculation, it would theoretically result in an adaptive over dependence on type 2 fibres for force generation, with faster muscle contractions but more rapid fatigue. A similar change in fibre type is recognized in mitochondrial myopathy, where histopathology is reported to show an increased ratio of type 2 to type 1 fibres^34^. More recently, proteins involved in mitochondrial fusion/fission have been implicated in the intracellular signalling processes regulating fibre type switching^35^, as observed in response to exercise^36^.

Although our biological metrics returned to levels that were compatible with controls and reported reduced levels of fatigue (Figure 1B), we found no significant correlation between changes in these two. This is perhaps not surprising. While chronic fatigue will place similar limitations on all, how a sufferer copes with this - and thus assesses the impact on their life - will also be affected by psychological factors such as level of resilience or access to support systems. The magnitude of change in FIS score, measured by a subjective questionnaire, is likely to be influenced at least as much by these factors as the underlying pathology. Our findings have significant, but indirect, clinical implications. Understanding the trajectory of neurophysiological recovery in long COVID has the potential to inform personalized treatment approaches and rehabilitation strategies. In particular, the measurement of peripheral fatigue is simple and can be achieved with low-cost equipment. This could deliver an objective assessment, to be interpreted alongside subjective measures such as the FIS score.

## Acknowledgements

The authors would like to thank the participants for their time and Norman Charlton for mechanical engineering support.

## Author Contributions

DSS, MRB, and SNB designed the study. NJM and AMEB collected the data. NJM processed the data. NJM, DSS, and MG performed analyses. NJM and MG wrote the manuscript. All authors edited and approved the final manuscript.

## Data Availability Statement

A spreadsheet containing values for all subject measurements across visits is available in Supplementary Material.

